# Adaptive hearing aid benefit in children with mild/moderate hearing loss: A registered, double-blind, randomized clinical trial

**DOI:** 10.1101/2021.07.14.21260541

**Authors:** Hannah J. Stewart, Erin K. Cash, Joseph Pinkl, Cecilia Nakeva von Mentzer, CCHMC Division of Audiology, Lisa L. Hunter, David R. Moore

## Abstract

**Objectives:** We completed a registered double-blind randomized control trial to compare acclimatization to two hearing aid algorithms by experienced pediatric hearing aid users with mild to moderate hearing loss. We hypothesized that extended use (up to 13 months) of the adaptive algorithm with integrated directionality and noise reduction, OpenSound Navigator (OSN), would result in improved performance compared to the control algorithm, omnidirectional (Omni), on auditory, cognitive and academic tasks.

**Design:** We recruited 42 children, aged 6 - 13 years old, through Cincinnati Children’s Hospital Medical Center’s Division of Audiology. Two children withdrew from the study due to noncompliance or discomfort. The remaining 40 children were paired by age (within one year) and hearing loss (level and configuration). The individuals from each pair were randomly assigned to a group: OSN (experimental) and Omni (control). Each child completed an audiology evaluation, hearing aid fitting, a follow up audiological appointment and two research visits up to 13 months apart. Research visit outcome measures covered speech perception (in quiet and in noise), novel grammar and word learning, cognition, academic ability and caregiver report of listening behaviours.

**Results:** The children with the experimental algorithm did not show improved performance on the outcome measures compared to the control algorithm. There was a significant relationship between age at first hearing aid use and Reading and Mathematical ability across all participants. Data from six children were not included in the analysis due to daily hearing aid usage of less than 6 hours.

**Conclusions:** Use of the experimental algorithm (OSN) neither enhanced nor reduced performance on auditory, cognitive and academic tasks compared to the control algorithm (Omni).

## INTRODUCTION

About 15% of children aged 6-19 years in the US have pure tone average (PTA) hearing thresholds > 15 dB HL (Niskar et al., 1998; Su & Chan, 2017), the level at which hearing loss in children is considered functionally significant. Typically, these children have bilateral PTA ≥ 26 dB HL, the threshold used in Butcher and colleagues’ (2019) meta-analysis to calculate a 1.1 per 1000 prevalence of bilateral hearing loss at newborn screening. Both children (Tomblin et al., 2014) and adults (Gatehouse, 1992) have been shown to obtain greater benefit from hearing aid use across time following initial fitting (acclimatization). While such acclimatization is controversial in adults, with several studies finding no greater, long-term benefit (Bender et al., 1993; Dawes et al., 2014; Humes et al., 2002; Humes & Wilson, 2003; Taylor, 1993), the weight of evidence in children favors increased benefit (Moeller & Tomblin, 2015). This issue is clinically significant because some of the same evidence shows the importance of earliest possible intervention for hearing loss (Moeller & Tomblin, 2015). The greater improvement in language and academic skills obtained by early hearing aid use presumably reflects improved speech perception through a process of auditory perceptual learning (Walker et al., 2020).

Intervention for hearing impairment is especially important in children who are learning language through everyday activities and in educational settings (McCreery et al., 2019; Walker et al., 2015a, 2020). Hearing aids are ideally used extensively during waking hours (Muñoz & Hill, 2015), and provide enhanced speech intelligibility (Ertmer, 2011), enabling long duration exposure to socially- and educationally-relevant stimulation. However, children wearing hearing aids exhibit greater listening effort evidenced by slower reaction times to verbal instructions, especially at more adverse signal to noise ratios, which may negatively impact upon learning and academic achievement in the classroom (McGarrigle et al., 2019). Both duration and salience of stimulation are considered to be key elements of perceptual learning (Wildt et al., 2019). Children also learn more easily than adults (Lucas et al., 2014), possibly due to enhanced brain plasticity and/or greater motivation with the task regimen.

It seems plausible that improving the features of hearing aids that are most useful for children, such as directional microphones (Chung & Zeng, 2009; Gravel et al., 1999), may further improve acclimatization. For example, children may be more dependent than adults on spatial localization of sound, due to reduced selective attention (Wightman et al., 2006), and the need to learn in noisy classrooms (Picard & Bradley, 2001) and home environments (Erickson & Newman, 2017). In response to these learning and environmental considerations, OpenSound Navigator (OSN) was developed to provide adaptive, integrated directionality and noise reduction. Briefly, OSN utilizes a dual microphone array to phasically analyze the acoustic environment. Subsequent directionality and noise reduction processing attenuate noise sources and diffuse noise respectively. The system updates 500 times/second across 16 independent frequency channels, enabling a rapid, spatial-based adaptive system with high selectivity to speech sounds (Le Goff et al., 2016).

In a previous, exploratory study (Pinkl et al., 2021), we showed that 6-12 year old children fitted with OSN-enabled hearing aids for two months received enhanced caregiver assessments of their speech and sound perception, spatial sound awareness and ability to participate in conversations. However, their measured speech perception, language, cognitive and academic skills were unaffected by using the hearing aids. As there was no control group in that study, it is possible that the enhanced assessments were affected by reporter bias. In addition, the acclimatization period was short and the sample size limited. Finally, it is possible that the range of measures used, although wide, missed other important aspects of listening and learning.

Browning et al. (2019) assessed speech in noise perception in children with OSN or omnidirectional (Omni) programming after no acclimatisation. They found that speech in steady state noise (SSN) thresholds were better for OSN than for Omni programming, regardless of whether the target talker was facing the participant or not. No programming difference was found for a speech in two-talker speech condition. However, they used a within-subject design for hearing aid programming in that the children received both programming options for testing rather than assessing a separate Omni control group.

To address these gaps in evidence, we designed a new, expanded, registered double-blind randomized control trial that compared extended (up to 13 months) use of the OSN algorithm with a control, Omni algorithm, programmed into the physically identical Oticon OPN hearing aids. We added outcome measures for speech in noise (BKB-SiN), statistical grammar learning and participant fatigue to our previous behavioural testing battery of speech perception, cognition, academic and caregiver report outcomes. We hypothesized that extended use of the OSN algorithm would result in improved performance on the range of skills previously examined and on other skills (statistical learning, and self-report effort measures) newly introduced in this study.

## MATERIALS AND METHODS

### Participants

#### Recruitment

Eligible participants were initially identified through a Cincinnati Children’s Hospital Medical Center (CCHMC) Division of Audiology medical record search. Study staff pre-screened each eligible child and mailed paper recruitment materials. After 2 weeks, families were contacted by phone and/or email regarding their interest in the study. If interested, the parent/guardian of the potential participant completed a series of online questionnaires. If all eligibility criteria (see below) were met, study staff consented the parent/guardian over the phone using the eConsent Framework in the Research Electronic Data Capture (REDCap; Harris et al., 2009) platform, also used for collecting and managing all study data. REDCap is a secure, web-based application providing: 1) an intuitive interface for validated data entry; 2) audit trails for tracking data manipulation and export procedures; 3) automated export procedures for seamless data downloads to common statistical packages; and 4) procedures for importing data from external sources. Participants were assented if they were 11 years of age or older.

#### Inclusion

Forty-two children were enrolled, but two were withdrawn due to noncompliance or discomfort. The 40 remaining were experienced pediatric HA users, ages 6,7 to 13,2 (years,months, mean = 9,9, Fig. 1). Age of hearing loss diagnosis ranged from birth to 8 years (mean = 3,8; Table 1), while age at receipt of first hearing aids ranged from 3 months to 9 years (mean = 4,1). Inclusion criteria were a) native English speakers, b) no history of ear surgeries, c) symmetrical sensorineural hearing loss in the mild to moderately-severe range from 500-4000 Hz (see “Audiological evaluation” below), d) no history of developmental delays, e) no medical diagnoses of neurologic/psychiatric disorders or attention deficits, f) history of consistent binaural HA use, and g) no prior experience with Oticon’s OSN algorithm. A further criterion was magnetic resonance imaging (MRI) compatibility (MRI results to be reported elsewhere). Three participants who were not MRI-compatible were enrolled to provide matches for other participants (one of whom was withdrawn).

**Table 1:** Participant demographics and summary of hearing aid models used prior to the study along with how long they wore their study hearing aids (time from HA fitting to RV2).

| Pair | Group | Shape of HL | R PTA | L PTA | Maternal education | Age at | Age of | Age at | Previous HA Type |  | Time with | Time with | RV2 during<br>COVID<br>pandemic |
| --- | --- | --- | --- | --- | --- | --- | --- | --- | --- | --- | --- | --- | --- |
|  |  |  |  |  |  | RV1<br>(y, m) | diagnosis<br>(y) | first HA<br>(y) | Make | Model | (y, m) | (m) |  |
| 1 | OSN | Bowl/notch | 40.0 | 53.3 | No high school | 8, 7 | 6 | 6 | Oticon | Sensei Pro | 2, 0 | 8 | no |
|  | omni | Bowl/notch | 43.3 | 43.3 | College degree | 9, 1 | 1 | 5 | Widex | Dream 220 | 3, 2 | 8 | no |
| 2 | OSN | Bowl/notch | 71.7 | 76.7 | College degree | 11, 6 | 1 | 1 | Phonak | Sky Q-70 M13 | 4, 4 | 11 | no |
|  | omni | Bowl/notch | 58.3 | 66.7 | College degree | 13, 4 | 2 | 2 | Phonak | Sky V50 P | 2, 9 | 6 | no |
| 3 | OSN | Falling | 45.0 | 36.7 | College degree | 7, 7 | birth | 9 months | Oticon | Sensei Pro | 1, 8 | 8 | no |
|  | omni | Falling | 61.7 | 63.3 | College degree | 8, 5 | 5 | 5 | Oticon | Sensei Pro SP | 1, 7 | 8 | no |
| 4 | OSN | Falling | 20.0 | 20.0 | College degree | 11, 5 | 3 | 3 | Oticon | Safari 600P | 8, 4 | 8 | no |
|  | omni | Falling | 23.3 | 25.0 | College degree | 11, 7 | 1 | 1 | Phonak | Sky Q50 M13 | 4, 5 | 9 | no |
| 5 | OSN | Falling | 23.3 | 26.7 | Some college | 11, 10 | 8 | 8 | Phonak | Sky V90 M | 2, 2 | 7 | no |
|  | omni | Bowl/notch | 50.0 | 46.7 | No high school | 11, 8 | 7 | 8 | Phonak | Sky Q-50 SP | 3, 11 | 8 | no |
| 6 | OSN | Flat | 68.3 | 68.3 | College degree | 11, 0 | 3 | 3 | Phonak | Sky Q50-SP | 4, 7 | 9 | no |
|  | omni | Flat | 65.0 | 61.7 | Post-graduate degree | 10, 0 | birth | 2 months | Phonak | Bolero V 90-P | 3, 3 | 11 | no |
| 7 | OSN | Rising | 35.0 | 36.7 | Some college | 10, 6 | 3 | 3 | Oticon | Safari 600P | 6, 2 | 10 | no |
|  | omni | Rising | 31.7 | 31.7 | Post-graduate degree | 9, 6 | 5 | 6 | Phonak | Sky Q50-M13 | 2, 7 | 9 | no |
| 8 | OSN | Bowl/notch | 28.3 | 36.7 | Some college | 9, 9 | 5 | 5 | Phonak | Sky 50 Q M 13 | 4, 6 | 12 | no |
|  | omni | Falling | 30.0 | 23.3 | College degree | 10, 0 | 1 | 1 | Phonak | Nios S H20 III | 5, 5 | 8 | no |
| 9 | OSN | Bowl/notch | 40.0 | 41.7 | College degree | 6, 8 | 4 | 4 | Oticon | Sensei Pro | 2, 0 | 11 | no |
|  | omni | Bowl/notch | 43.3 | 45.0 | College degree | 6, 2 | 4 months | 9 months | Oticon | Safari 600P | 5, 6 | 10 | no |
| 10 | OSN | Falling | 41.7 | 41.7 | College degree | 11, 7 | 0 | 3 | Widex | Inteo BTE | 7, 11 | 8 | no |
|  | omni | Bowl/notch | 31.7 | 38.3 | Some college | 12, 4 | 3 | 3 | Oticon | Safari | 8, 0 | 9 | no |
| 11 | OSN | Rising | 26.7 | 30.0 | College degree | 8, 10 | 6 | 6 | Oticon | Sensei Pro | 2, 2 | 15 | yes |
|  | omni | Falling | 40.0 | 23.3 | College degree | 9, 8 | 7 | 7 | Oticon | Sensei Pro | 2, 6 | 9 | no |
| 12 | OSN | Bowl/notch | 51.7 | 51.7 | College degree | 11, 11 | 3-6 months | 3-6 months | Oticon | Sensei Pro | 2, 9 | 8 | yes |
|  | omni | Bowl/notch | 56.7 | 45.0 | Some college | 10, 10 | 8 | 8 | Phonak | Sky Q 50 SP | 2, 7 | 12 | yes |
| 13 | OSN | Flat | 45.0 | 45.0 | Some college | 7, 6 | 5 | 5 | Oticon | Sensei Pro | 1, 11 | 9 | yes |
|  | omni | Rising | 38.3 | 43.3 | Some college | 7, 11 | 1 month | 2 months | Phonak | Sky Q50 SP | 3, 8 | 10 | no |
| 14 | OSN | Flat | 48.3 | 48.3 | College degree | 8, 6 | 1 month | 5 months | Oticon | Safari 600 P | 8, 1 | 13 | yes |
|  | omni | Flat | 48.3 | 48.3 | Some college | 8, 7 | 4 | 4 | Widex | Dream 440 | 4, 7 | 13 | yes |
| 15 | OSN | Falling | 51.7 | 50.0 | Some college | 10, 5 | 4 months | 5 months | Oticon | Vigo Pro | 9, 11 | 9 | no |
|  | omni | Falling | 10.0 | 11.7 | College degree | 9, 4 | 5 | 6 | Phonak | Sky Q 50 M13 | 3, 3 | 10 | no |
| 16 | OSN | Bowl/notch | 58.3 | 58.3 | College degree | 9, 5 | 4 | 5 | Phonak | Sky Q70 SP | 4, 5 | 10 | no |
|  | omni | Flat | 30.0 | 30.0 | Post-graduate degree | 9, 7 | Unknown | Unknown | Unknown | Unknown | 9, 4 | 9 | no |
| 17 | OSN | Flat | 36.7 | 38.3 | Some high school | 10, 11 | 8 | 9 | Oticon | Sensei Pro | 1, 10 | 9 | no |
|  | omni | Rising | 43.3 | 50.0 | Some college | 11, 11 | 7 | 7 | Resound | LiNX3D977 | 1, 10 | 10 | yes |
| 18 | OSN | Flat | 35.0 | 35.0 | Post-graduate degree | 10, 4 | 4 | 4 | Oticon | Safari 600 | 5, 10 | 13 | yes |
|  | omni | Falling | 35.0 | 36.7 | Post-graduate degree | 10, 5 | 5 | 5 | Oticon | Sensei Pro | 1, 1 | 11 | no |
| 19 | OSN | Flat | 45.0 | 48.3 | Post-graduate degree | 6, 10 | 2 | 2 | Phonak | Sky Q50 m13 | 4, 2 | 9 | no |
|  | omni | Flat | 45.0 | 45.0 | Some college | 7, 5 | 1 month | 3 months | Phonak | SkyQ 50 m13 | 3, 5 | 7 | no |
| 20 | OSN | Falling | 36.7 | 35.0 | Post-graduate degree | 9, 7 | 5 | 5 | Oticon | Sensei Pro | 5, 0 | 11 | no |
|  | omni | Falling | 31.7 | 31.7 | College degree | 10, 2 | 2 | 2 | Phonak | Sky V90-P | 2, 2 | 8 | no |

**Figure 1:**
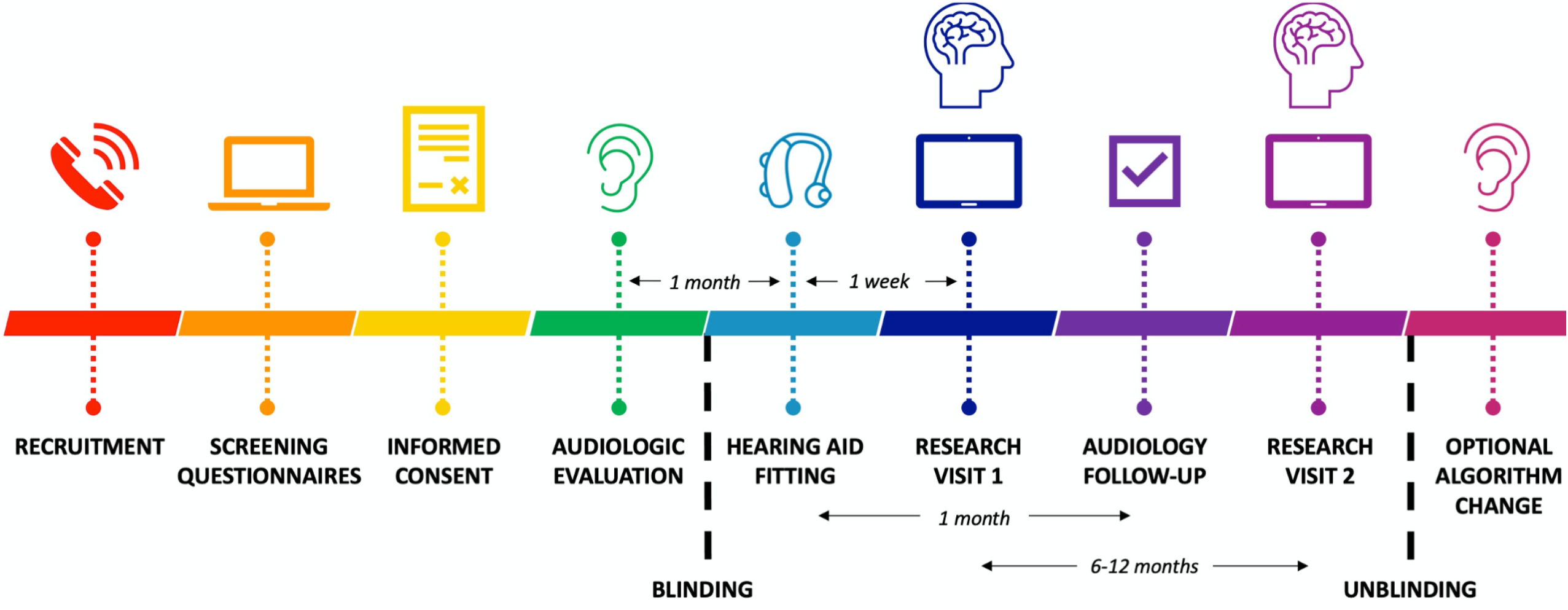
Study timeline.

#### Ethics and preregistration

Ethical approval for all clinical tests, services and data collection procedures was obtained from the CCHMC Institutional Review Board. The study was preregistered with clinicaltrials.gov (NCT03771287).

#### Blinding and Randomization

Participants were recruited in age (within one year) and hearing loss (level and configuration) matched pairs. The individuals in each pair were randomly assigned to a group; one child used the omnidirectional setting (control), and the other used the OpenSound Navigator (experimental). This randomization was assigned via a coin flip by a research coordinator who was not involved in recruitment, testing, or analysis for this study. Each child’s hearing aid algorithm was saved in REDCap; only accessible to the research coordinator who randomized the study pairs and the study audiologists. When research study staff scheduled each participant’s hearing aid fitting appointment, the external research coordinator notified the fitting audiologist. As part of standard of care, the hearing aid algorithm was enabled (OSN or omnidirectional) and added in the participants’ medical record notes. Participants, their caregivers, and study research staff were unaware of each participant’s enabled feature until both participants in any respective pair had completed the study.

#### Incentives

Participants received a payment of $30 for completing the first research and MRI visits and $60 for completing the second research and MRI visits. At the end of the study they received a $40 bonus if they wore their hearing aids for at least 8 hours a day (as per their logging data). The children were allowed to keep their study hearing aids and accessories at the end of the study.

### Procedures

The study schedule of participant visits is shown in Fig. 2. Many of the procedures have been described in detail previously (Pinkl et al., 2021) and will be presented only briefly here. All audiological visits followed a checklist of procedures to ensure that all study audiologists followed the same evaluation and fit protocols.

**Figure 2:**
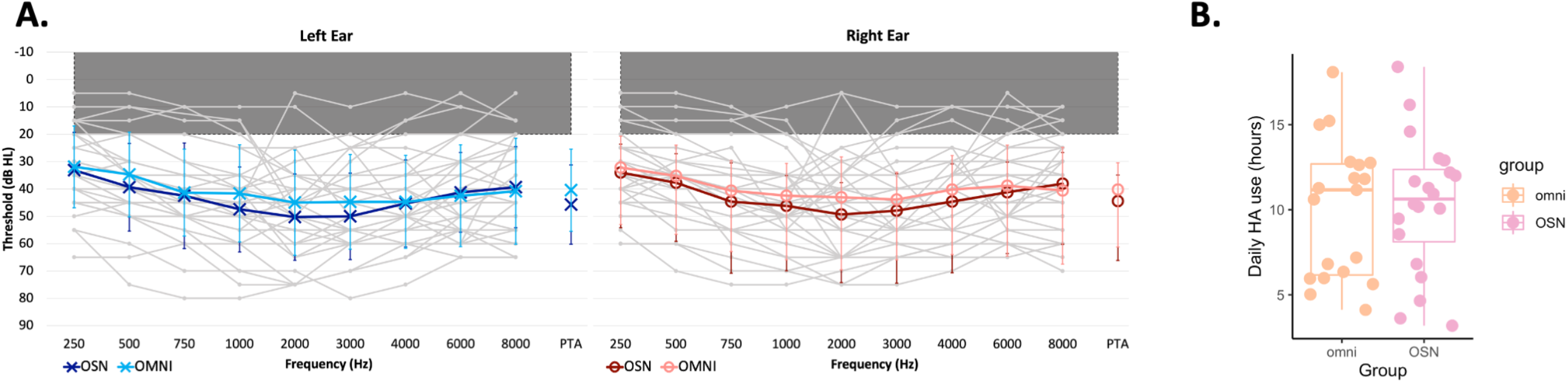
Participant details. (A) Audiograms showing individual thresholds in light grey and group means in color (OSN dark, OMNI light). PTAs calculated from 500, 1000 and 2000 Hz. (B) Average daily hearing aid usage during the study, as measured by the hearing aid logging feature. Boxplots show the groups’ upper quartile, median and lower quartile.

#### Audiologic evaluation

A clinical evaluation which included pure-tone thresholds obtained using the modified Hughson-Westlake procedure (Hughson and Westlake,1944), word recognition testing and immittance testing, was performed on all participants by licensed audiologists at the Division of Audiology, CCHMC. Air and bone conducted pure-tone signals were presented through EARTone 3A insert earphones and a MelMedtronics B71 adult bone oscillator, respectively, using a GSI 61 Clinical Audiometer (Grason-Stadler, Eden Prairie, MN). Pure-tone thresholds were obtained from 250-8000 Hz at half-octave increments via air-conduction and 500-4000 Hz at octave increments via bone-conduction (Fig. 1).

Monaural and binaural speech testing was completed via air-conduction and included speech reception thresholds (SRT) and monosyllabic word recognition. Speech recognition testing was performed using recorded male voices with open-set lists (NU-6, W-22 or PKB; Auditec, Inc.) presented in quiet at 40 dB SL (sensation level; based on SRT) or the participant’s most-comfortable level.

Standard 226-Hz tympanometry (Hunter & Blankenship, 2017) was completed using a GSI Tympstar Middle Ear Analyzer or Titan/IMP440. Previous audiometric results were used for participants who received an audiologic evaluation at CCHMC within six months of study enrollment. Earmold impressions were also taken at this visit; participants were permitted to continue use of prior earmolds if the audiologists deemed they provided adequate fit and comfort.

#### Hearing aid fitting

Within one month of the audiologic evaluation, bilateral Oticon OPN 1 PP behind-the-ear (BTE) HA’s with standard tubing, custom ear molds and a compatible wireless microphone and Bluetooth streamer (ConnectClip), were dispensed for each participant. Ear molds were either skeleton or canal style and made from silicone material with venting size selected by the fitting audiologist. Manual controls on the hearing aids were disabled. The OSN algorithm was enabled, where used (see Randomization, above), with the transition and noise reduction in simple/complex environments set to the manufacturer default setting.

HA verifications were performed using Verifit 2.0 (Audioscan, Dorchester, ON, Canada). To measure the Speech Intelligibility Index (SII) real-ear speech mapping was completed using a standard recorded passage presented at 55, 65 and 75 dB (low, average and loud, respectively) sound pressure level (SPL). Prescriptive targets were set using Desired Sensation Level (DSL) v5.0 based on each participant’s audiometric thresholds and individual real ear-to-coupler differences. Real-ear probe microphone measurements were used to ensure that HA gain met prescriptive targets. Fine-tuning gain adjustments were made so that HA output was within 5 dB at 250, 500, 1000 and 2000 Hz and 8 dB at 3000 and 4000 Hz of the prescriptive targets (Bagatto et al., 2016; Bagatto et al., 2011). Participants and accompanying caregivers were instructed on HA use and care by the fitting audiologist. They were asked to ensure hearing aids were used whenever possible, with a minimum of 8 hours per day. Participants that utilized hearing aid compatible assistive listening devices and bluetooth streaming for classroom learning were instructed to continue using those same devices with adapters and audio boots provided by Oticon when necessary.

#### Research visit 1 (RV1)

Participants completed the first round of experimental behavioral testing, as well as an initial MRI, 2-7 days after hearing aid fitting. The test battery included 6 different assessments: a free-field word repetition task, sentence repetition tasks in quiet and in noise, a novel word learning task, a set of standardized cognition tasks, and an academic achievement task. Parents of participants were asked to complete caregiver report questionnaires that assessed their child’s speech and hearing abilities.

### Audiology follow-up

One month after hearing aid fitting, the fitting audiologist checked the hearing aid fit and advised on the child’s average daily usage. Validation measures included aided free-field narrow-band thresholds (center frequencies of 500, 1000, 2000 and 4000 Hz) and speech recognition in quiet testing (NU-6, W-22 or PBK lists, consistent with the initial audiologic test). Word recognition was completed using recorded male voices presented at 35 and 55 dB HL presented at 0° azimuth.

#### Research visit 2 (RV2)

Participants returned 6-13 months after their hearing aid fitting. The initial battery of testing was repeated, with different versions to reduce learning effects, in addition to a novel statistical grammar learning task. Caregivers completed the same questionnaires, this time in reference to their child’s speech and language abilities with their study hearing aids. A novel pediatric fatigue questionnaire, the PROMIS, was administered both as a self-report by the participant and, independently, as a caregiver report.

#### Optional Algorithm Change

Upon completion of the study, participants were unblinded and had the option of scheduling a final study follow-up appointment with their fitting audiologist for programming changes. At this appointment, they could change the hearing aid programming following discussion with the audiologist. Most participants chose to continue use of the OSN program, or to enable OSN if they had been in the OMNI group. Each participant’s daily hearing aid usage (Fig. 2B) was reviewed and discussed with caregivers.

#### Unblinding

Research staff involved in testing and/or analysis were unblinded after all participants had completed the study.

### Behavioral testing

All tests were performed in a double-walled sound-treated booth (IAC) with an observation window. Participants wore their study hearing aids in the experimental setting. Unless otherwise stated, auditory test stimuli were generated in Matlab and presented through an M-Track Eight interface (M-Audio, Cumberland, RI), Servo 120a power amplifiers (Samson Technologies, Hicksville, NY), and JBL Control 1Xtreme 4” loudspeakers in manufacturer’s enclosures (Harman International Industries, Stamford, CT), atop microphone stands at a height of 96 cm. The tester was outside the booth and communicated with the participant visually and via intercom.

Order of testing was randomized in a latin square design across participants. All tests were completed twice: within one week of hearing aid fitting (RV1; Fig. 2) and 6-13 months post-fitting (RV2). Test sessions of sentence repetition and novel word learning were recorded using Audacity (v. 2.3.2). Recordings were re-scored by a second researcher. If there was agreement between the two researchers, the initial score was used. Disagreements were reconciled by a third researcher.

#### Word repetition in noise

This test (Figure 3A) was adapted from custom software supplied by Boys Town National Research Hospital (Browning et al., 2019) and measured frontally-presented monosyllabic target word thresholds in rearward masking noise. Target words were from English reading lists suitable for 5 and 6 year olds (Corbin et al., 2016), normalized to a fixed intensity of 65 dBrms SPL and spoken by adult males in an American English dialect. Two-talker masker (TTM) speech streams were created from separate passages from *Jack and the Beanstalk*. Speech-shaped noise (SSN) was based on the spectral envelope of the TTM speech streams. Maskers were set at an initial level of 55 dBrms SPL. Three test conditions varied either the target direction or the noise type (Fig. 3A).

**Figure 3:**
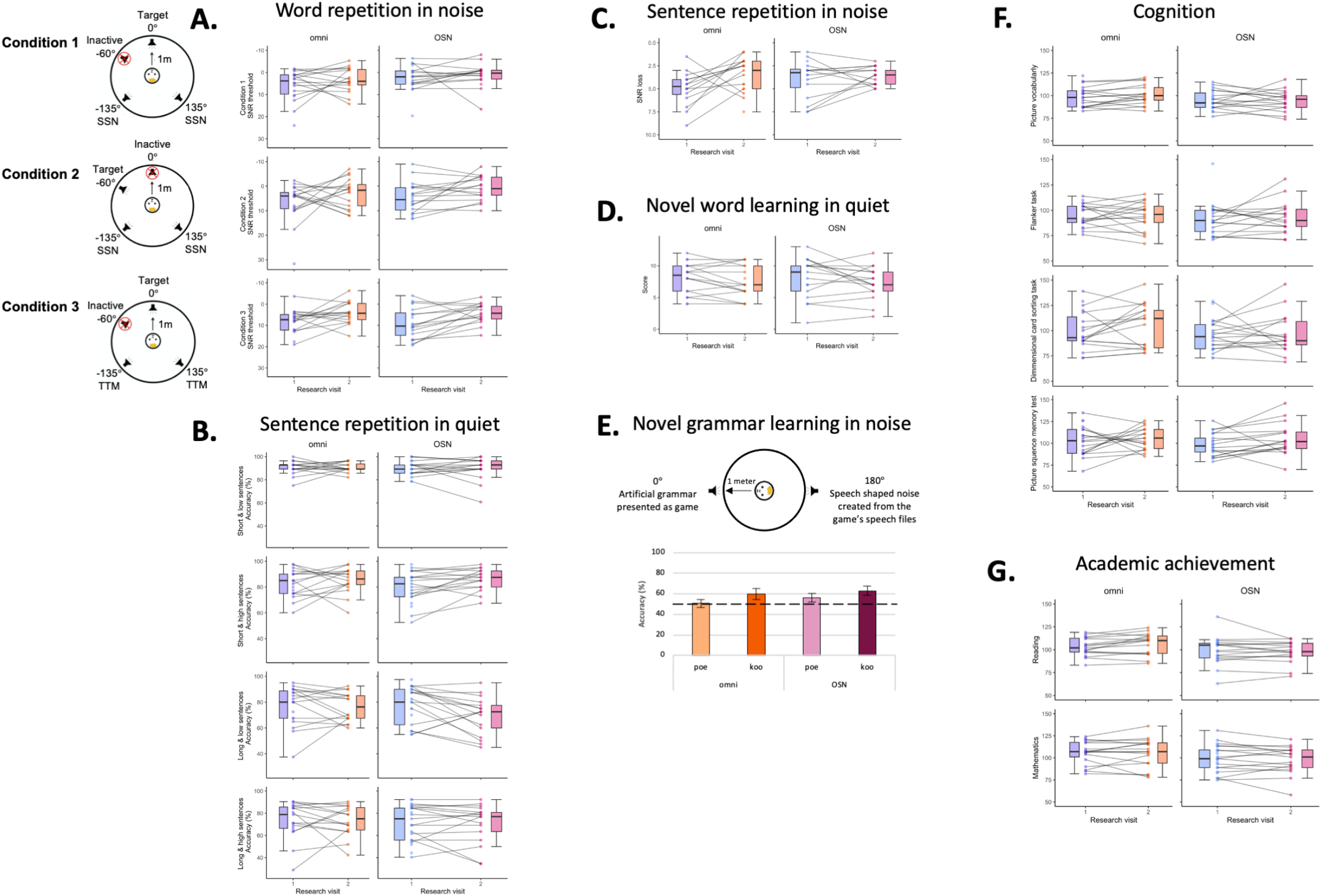
Behavioral test performance. No difference was found between the Omni and OSN group on any task. (A) Word repetition in noise (Browning et al., 2019) signal-to-noise ratio (SNR) thresholds for words in noise for: Condition 1: speech presented at 0° azimuth with speech-shaped noise (SSN) maskers behind the participant at ±135°; Condition 2: speech presented at 60° azimuth with SSN maskers presented at ±135°; Condition 3: speech presented at 0° azimuth with two-talker speech masking (TTM) maskers presented at ±135°. (B) Sentence repetition in quiet (Moll et al., 2015) scores for the four sentence types. (C) Sentence repetition in noise (Etymōtic Research, 2005). (D) Novel word learning in quiet showing standardized scores from the NEPSY (Brooks et al., 2009; Korkman et al., 1997), showing age standardized scores. (E) Novel grammar learning in noise, measured at RV2 only. The artificial grammar was presented at 0° azimuth with SSN presented at 180°. The horizontal dashed line on the graph marks vocabulary learning at chance, 50%. Error bars show SEM. (F) Cognition as measured by 4 subtasks from the NIH toolbox (Weintraub et al., 2013), showing age standardized scores: picture vocabulary; attention – flanker task; executive function – dimensional change card sorting task; episodic memory – picture sequence memory test. (G) Academic achievement as measured by two composite scores (reading and mathematics) from the Woodcock-Johnson IV Tests of Achievement (Schrank et al., 2014), showing age standardized scores. For all figures, the colored dots show individual scores with the lines between marking the change in the individual’s performance between RV 1 and 2. The boxplots show the groups 25th, 50th and 75th percentiles of the group with the whiskers indicating the groups maximum and minimum scores.

Participants used a chin rest for head and hearing aid microphone positioning to remain in the center of a speaker ring, diameter 2m. They were instructed to remain still, look straight ahead, listen closely for the target word, and repeat it back. If they were unsure what word they heard, they were instructed to guess. Correct/incorrect identification of Target increased/decreased masker level by 4 dB on next trial. After the second reversal, step size was reduced to 2 dB. Speech reception threshold (SRT) was the mean of the last 6 reversals, following 8 total reversals.

#### Sentence repetition in quiet

Speech perception, verbal working memory and grammatical knowledge were assessed by listening to and repeating recorded sentences (after Moll et al., 2015). The sentences were provided by an online research platform from Uppsala University (*Audio Research*, 2018; Ranjbar & Nakeva von Mentzer, 2020) and presented by a native female speaker at 50 dBrms SPL in Standard American English. Two loudspeakers were situated either side of a computer monitor (±5° azimuth) on a table 84 cm in front of the seated participant, who was instructed to listen closely to the stimuli, and repeat back the sentence. If the participant forgot or did not hear the sentence, they were encouraged to guess. Each sentence was scored based on the accuracy of word content and order. Sentences were categorized for memory by length (involving use or non-use of adjectives) and for grammar by complexity (use or nonuse of a ditransitive passive sentence structure where subject and object were expressed by two prepositions). Sentences could thus be short with low complexity (e.g. “The mom baked her daughter a pie.”), short with high complexity (e.g. “A piano was delivered by the dad to his son.”), long with low complexity (e.g. “The kind man ordered the tired woman a hot coffee.”), or long with high complexity (e.g. “A purple pencil was offered by a friendly girl to the new boy.”). Different sets of sixteen sentences comprised the task list at each research visit. Each set had four sentences from each of the four categories.

#### Sentence repetition in noise

The Bamford-Kowal-Bench Sentences in Noise (BKB-SiN) test contains 18 list pairs of recorded sentences in four-talker babble noise (Bench et al., 1979; Etymōtic Research, 2005). In this study, participants were presented with one list pair (2 × 10 sentences) at a constant 70 dBrms SPL from a frontal speaker. Sentences were preceded by the verbal cue “Ready” and spoken by a male voice in Standard American English. The concurrent babble noise level increased by 3 dB after each sentence, progressively reducing signal-to-noise ratio (SNR) from +21 to -6 dB across each 10 sentence sublist (Etymōtic Research, 2005; Holder et al., 2018). Participants were asked to repeat each sentence verbally and were scored by the number of correctly identified keywords, averaged across the two sublists. Reported scores are the SNR-50, the SNR at which 50% of target words were correctly identified, normalized as SNR loss relative to normal hearing listeners.

#### Novel word learning in quiet

The NEPSY-II, a standardized neuropsychological test battery, assesses multiple neurocognitive domains (Brooks et al., 2009; Korkman et al., 1997). The Repetition of Nonsense Words subtest gauges novel word-learning ability. Successful completion of the task requires participants to decode phonological stimuli and articulate novel words. Thirteen recorded nonsensical words in a male voice were presented at 65 dB SPL through the same speaker configuration as the sentence repetition in quiet task. Words varied from two to five syllables in length, where one correctly-pronounced syllable amounted to one point. Points from each word were summed to formulate a composite score, which was then scaled by age.

#### Novel grammar learning in noise

The children were exposed to a novel grammar (von Koss Torkildsen et al., 2013) in the form of a game where they were teaching an alien a new language. During a single exposure at RV2, children were introduced to an aX and Yb grammar, with a and b components consisting of a single syllable nonword (CV), X representing one-syllable nonwords (CVC), and Y representing one-syllable nonwords (CVCCV). Examples of the aX ‘poe’ grammar are “poe zek” and “poe zug”, whereas Yb ‘koo’ grammar is exemplified by “wagso koo” and “zikvoe koo”. After four examples, to introduce the participant to the format of the task, the grammar exposure was split into 12 blocks, each 4 trials long ending with 2 two-alternative forced choice trials to assess their learning. This totaled 48 exposures to the grammar with 24 test trials to assess grammar knowledge. Target words were presented at 0° azimuth and speech shaped background noise, created from the spectral envelope of the nonwords used in the task, was presented at 180° azimuth throughout the task.

#### Cognition

The NIH Toolbox Cognition Battery assesses the brain’s higher-level functions language, perception, planning and execution of behavior, and memory (Weintraub et al., 2013). Four subtests from this battery, each lasting 5-15 minutes, were administered to participants on an iPad with the tester seated next to them. Individual subtests produced a raw score as well as an age-standardized score. Together the results of each standardized subtest comprise the ‘Early Childhood Composite’ score.

- The Picture Vocabulary Test assesses receptive vocabulary. Four pictures were presented together on the screen, and a single word was spoken by a female voice in Standard American English. Participants were instructed to select the picture that best matched the meaning of the spoken word.
- The Flanker Inhibitory Control and Attention Test measures attention and inhibitory control, an element of executive function. Five shapes (fish or arrows) were presented in a row on the screen, each pointing either left or right. Participants selected the direction (left or right) of the middle stimulus, while ignoring the distractors, as quickly as possible. Accuracy and response time were factored into scoring.
- The Dimensional Change Card Sort Test assesses the planning and execution of behavior (attention shifting, executive function). In each trial, participants were presented one of two shapes (e.g. a ball or a boat) that could be either yellow or blue. The word “shape” or “color” appeared on the screen, instructing the participant how to choose quickly the appropriate matching item.
- The Picture Sequence Memory Test assesses episodic memory. Illustrations were presented in a specific order with verbal narration, after which they were randomly shuffled. Participants were asked to place the illustrations back in the order in which they were presented.

#### Academic achievement

Four standardized subtests were selected from The Woodcock-Johnson IV (Schrank et al., 2014) to test reading and mathematical ability. Questions progressively increased in difficulty throughout each subtest with scoring procedures following basal and ceiling rules. Participants were guided through the assessment by the tester, who was seated next to them.

- The *Letter-Word Identification* test measures word identification ability and pronunciation accuracy. Single words were listed vertically on a screen, and the participant was instructed to read each word out loud from top to bottom.
- The *Passage Comprehension* test measures reading comprehension. Younger children are initially presented with pictures and must point to a specific one upon tester instruction. These transition to sentences with a word missing wherein the participant must silently read the sentence and tell the tester what the missing word is. Older children were tasked with lengthier passages that required the use of context clues to determine the missing word.
- The *Applied Problems* test presented math problems in word or picture form. Participants were given a pencil and scrap paper to use at their discretion.
- The *Calculation* test assesses straightforward mathematical knowledge. Participants were given a worksheet that began with simple numerical calculations (addition, subtraction, etc.) and eventually geometric, logarithmic, and calculus-based problems.

### Questionnaires

#### Speech, Spatial and Qualities of Hearing Scale (SSQ)

The SSQ (Gatehouse & Noble, 2004) is a self-report measure of hearing ability in varying contexts. This study used a version adapted for children (Galvin & Noble, 2013) in which caregivers rated their child’s listening ability in everyday scenarios on a scale from 0-100. Twenty-seven items were included across four categories: speech hearing, spatial hearing, qualities of hearing, and conversational uses of hearing. A higher average score for each category indicated better ability. This questionnaire was also completed at both RV1 and RV2 (Fig. 4).

**Figure 4:**
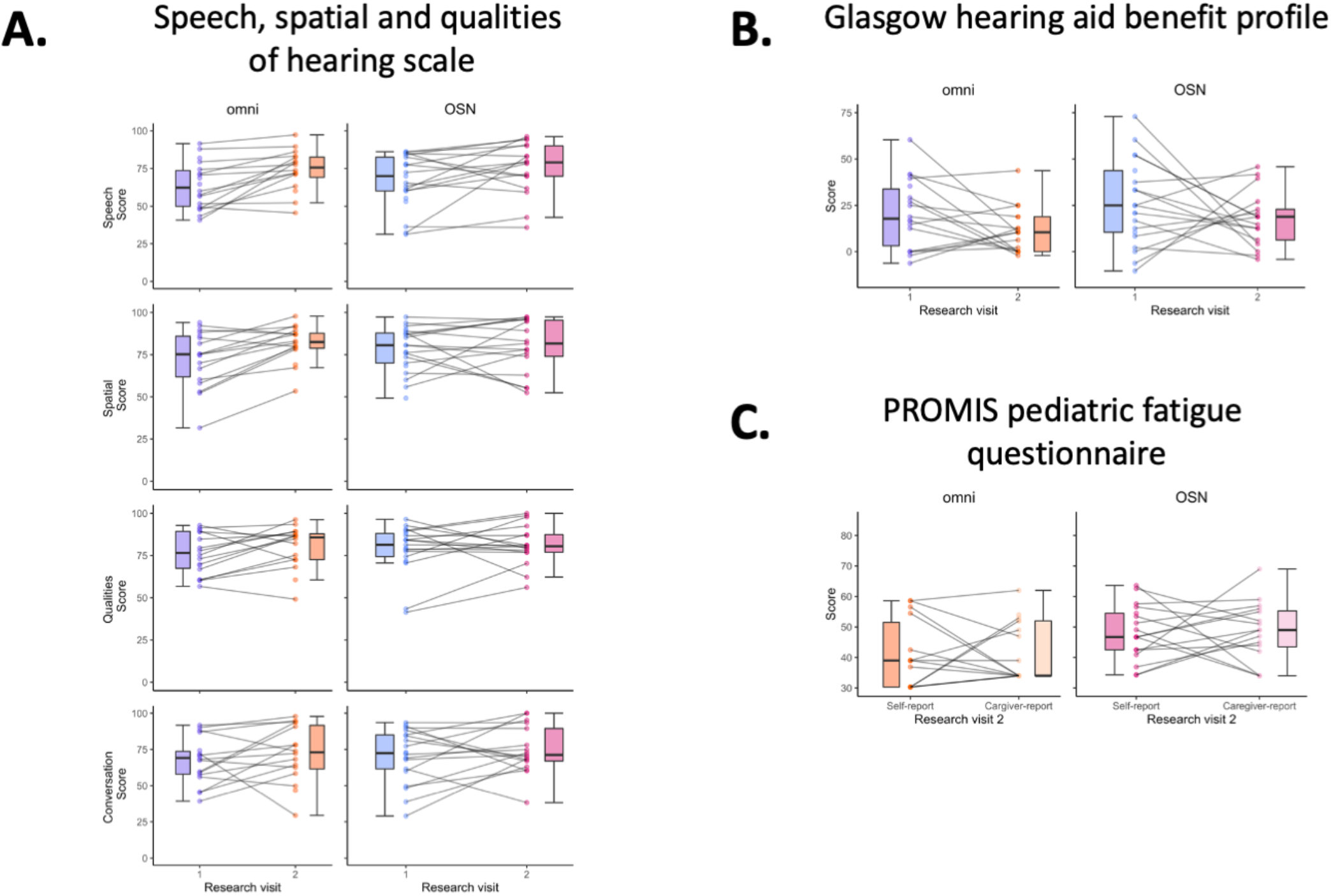
Questionnaires. (A) Speech, spatial and qualities of hearing scale (SSQ; Galvin & Noble, 2013). (B) Glasgow hearing aid benefit profile (Kubba et al., 2004). For figures (A) and (B), the colored dots show individual scores with the lines linking each individual’s performance between RV1 and RV2. (C) PROMIS pediatric fatigue questionnaire showing standardized T-scores from the self- and caregiver-reports (Varni et al., 2014). Lower scores indicate more frequently reported fatigue. Colored dots show individual scores and lines link self-with caregiver-report, both measured at RV 2 only. In all graphs the boxplots show group 25th, 50th and 75th percentiles and the whiskers indicate maximum and minimum scores.

#### Glasgow Hearing Aid Benefit Profile

The GHABP (Gatehouse, 1999) is a self-report measure of the effectiveness of assistive technology in hearing-impaired adults. In this study, a modified pediatric version based on caregiver report was used (Kubba et al., 2004). The Glasgow Children’s Benefit Inventory (GCBI) consists of 24 questions with subscales of hearing disability, handicap, hearing aid usage, hearing aid benefit, hearing aid satisfaction, and residual disability relative to the benefit of prior hearing aids. This questionnaire was administered at both RV1 and RV2. All questions utilized a five-point Likert scale with each scalepoint corresponding to a numerical score (100 = much better, 50 = a little better, 0 = no change, -50 = a little worse, -100 = much worse). The 24 responses were averaged to create a composite wherein higher/more positive values reflected greater intervention benefit.

#### Pediatric Fatigue Questionnaire

The Patient-Reported Outcomes Measurement Information System (PROMIS) short form includes measures of physical, mental, and social health (Irwin et al., 2010; Lai et al., 2013; Varni et al., 2014). The shortened form of the Pediatric Fatigue measure includes 10 items assessing feelings of tiredness on a five-point Likert scale (1 = Never, 2 = Almost Never, 3 = Sometimes, 4 = Often, 5 = Almost Always; Lai et al., 2013). Recent research suggests that hearing impairment may increase risk for fatigue due to the increased need for deliberate listening and attention (Hornsby et al., 2016). Though much of this evidence is based on subjective accounts, the use of patient-reported outcomes is arguably the most direct source of information on health outcomes (Gerhardt et al., 2018). Participants completed a self-report version with the assistance of study staff, and parents completed a proxy version. Scores were averaged to create a composite for each version; higher scores indicate more frequent feelings of fatigue. The PROMIS Pediatric Fatigue measure was administered only at RV2.

### Statistical analysis

Participants who logged less than 6 hours of daily HA use (OSN n = 3, omni n = 3) and an OSN participant with HA technical difficulties were not included in the analysis. Repeated-measures analyses of variance (ANOVAs) were used to assess performance differences between RV1 and RV2, and between the OSN and Omni groups. Greenhouse-Geisser correction was applied to variables that failed tests of normality. Post-hoc t-tests were Bonferroni-Holm corrected. Where available, standardized test scores were used. However, if a test did not provide standardized scores (word repetition in noise, sentence repetition in quiet and in noise, novel grammar learning in noise) age was included as a covariate.

Novel grammar learning in noise was administered only at RV2. Participants who showed a clear button preference (i.e. clicking the same button for the majority of the trials: OSN n = 3, omni n = 4) were excluded from the analysis. A repeated measures ANOVA compared accuracy on the aX and Yb grammars. Planned follow up analysis used one sample t-tests to compare the accuracies of these grammars to chance (50%). The PROMIS questionnaire was also only administered at RV2. One way ANOVAs were used separately for the self-report and the caregiver report scores. A planned paired samples t-test was used to compare if the self- and caregiver reports were significantly different from one another.

Pearson correlations were used to explore the relationship between speech perception, language, cognitive and academic skills with age of first hearing aid fit and daily hearing aid usage. P-values were adjusted for multiple comparisons whereby each test (e.g. word repetition in noise, NIH toolbox) was treated as its own “family”. Adjustments were made for the following number of comparisons: three for word repetition in noise (0.017), four for sentence repetition in quiet (0.0125), one for sentence repetition in noise (0.05), one for novel word learning in quiet (0.05), two for novel grammar learning in noise (0.025), four for the NIH toolbox assessing cognition (0.0125) and two for the WJ-IV assessing academic ability (0.025).

## RESULTS

### Audiometry

The two groups (OSN and OMNI) were well matched for age, mean hearing loss and interaural symmetry (Table 1; Fig. 2A) with no mean PTA group difference (left p = 0.27; right p = 0.38). However, there was a wide range of individual audiogram configurations and hearing loss severity.

### Behavioural testing

#### Word repetition in noise (Fig. 3A)

Word repetition was poorer in the two-talker speech masker (Condition 3) compared with the speech-shaped noise masker (Conditions 1; t = -3.85, p < 0.001, d = -0.69; and 2; t = -3.34, p = 0.003, d = -0.60) but was unaffected by whether the target was in front (Condition 1) or 60°left (Condition 2) of the listener (F_2,54_ = 4.68, p = 0.013, = 0.15). Performance improved across time between RV 1 and 2 (F_1,27_ = 6.24; p = 0.019, = 0.19). OSN use did not affect performance significantly, and there were no significant interactions between OSN use and masker type or RV.

#### Sentence repetition in quiet (Fig. 3B)

Short sentences were reproduced more accurately than long sentences (F_1,28_ = 15.61, p < 0.001, = 0.36) and sentences of low complexity were reproduced more accurately than those of high complexity (F_1,28_ = 22.80, p < 0.001, = 0.45). However, OSN use did not affect performance significantly, and there were no significant interactions between OSN use and sentence length, complexity or duration of hearing aid use.

#### Sentence repetition in noise (Fig. 3C)

SNR loss scores were mostly in the range 3-7 dB, described by the test manual as a “Mild SNR loss”, where 0-3 dB is “Normal” and 7-15 dB is “Moderate”. Scores did not improve between tests and there was no significant difference between OSN and Omni groups overall or between tests.

#### Novel word learning in quiet (Fig. 3D)

There was no significant change in scaled scores between RV1 and RV2 and no significant difference between OSN and Omni groups overall or between tests.

#### Novel grammar learning in noise (Fig. 3E)

This was assessed at RV2 only, as the stimuli and paradigm needed to be novel. The participants learnt the Yb grammar significantly better than the aX grammar (F_1,20_ = 8.96, p = 0.007, = 0.31). Further analysis showed that the participants did not learn the aX grammar, i.e. where the grammar key ‘poe’ was at the start of the word, significantly more than chance (50%) but they did learn the Yb grammar (t_22_ = 3.03, p = 0.006, d = 0.63), i.e. where the grammar key ‘koo’ was at the end of the word. There was no significant difference between OSN and Omni groups overall.

#### Cognition (Fig. 3F)

Age-standardized scores have a mean of 100 and a SD of 15. Participant scores were in the range of 80-120 for all of the four subtests. There was a significant difference between subtest scores (F_3,93_ = 5.29, p = 0.005, = 0.15 Greenhouse-Geisser corrected) with picture sequence memory test performance significantly better than that for the flanker task (t = -3.97, p < 0.001, d = -0.69). There was no significant change between RV1 and RV2 and no significant difference between OSN and Omni groups overall or between research visits.

#### Academic achievement (Fig. 3G)

Age-standardized scores ranged between 90 and 120 on both outcomes and, compared with most other tests and excepting a few outliers, were remarkably stable across time for each individual. There was no significant change between RV1 and RV2 and no significant difference between OSN and Omni groups overall or between research visits.

### Questionnaires

#### Speech, Spatial and Qualities of Hearing Scale (SSQ; Fig. 4A)

Between RV1 and RV2, Speech (F_1,29_ = 15.03, p = 0.0012, = 0.34) and Spatial (F_1,29_ = 4.86, p = 0.036, = 0.14) scores increased, the Quality score (F_1,29_ = 270.44, p < 0.001; = 0.90) decreased, and there was no change for the Conversation score. Overall, there were no significant differences between OSN and Omni groups. However, there was an interaction between RV and group for the Spatial score (F_1,29_ = 5.56, p = 0.025, = 0.16). The Omni group score increased from RV1 to RV2, but OSN did not. This suggested that caregivers perceived an increased benefit of the Omni strategy as a result of prolonged use of that strategy.

#### Glasgow Hearing Aid Benefit Profile (Fig. 4B)

There was no significant change in scores between RV1 and RV2 and no significant difference between OSN and Omni groups, overall or between tests.

#### Pediatric Fatigue Questionnaire (PROMIS; Fig. 4C

This was administered at RV2 only and showed no significant difference between OSN and Omni groups on either the participant self-report scores or those by their caregivers. There was no significant difference between participants’ self-report and their caregiver scores.

### Hearing aid use

Given the similarity between OSN and Omni performance, we combined both groups to evaluate the effect of hearing aid fitting and use patterns. Participants’ age at first hearing aid fit (Table 1) correlated significantly with their WJ-IV Reading (r = -0.42, p = 0.016) and Mathematics (r = -0.43, p = 0.012) outcomes at RV2, but not at RV1 (Reading: r = -0.36, p = 0.037, note that this does not survive family correction of 0.025; Mathematics: r = -0.34, p = 0.052; Fig. 5A). The improvement in the flanker task between RV1 and RV2 correlated significantly with their average daily hearing aid usage (r = 0.47, p = 0.006; Fig. 5B). These comparisons at RV2 remained significant after family correction and indicated that the overall duration of hearing aid use was associated with better academic and cognitive outcomes.

**Figure 5:**
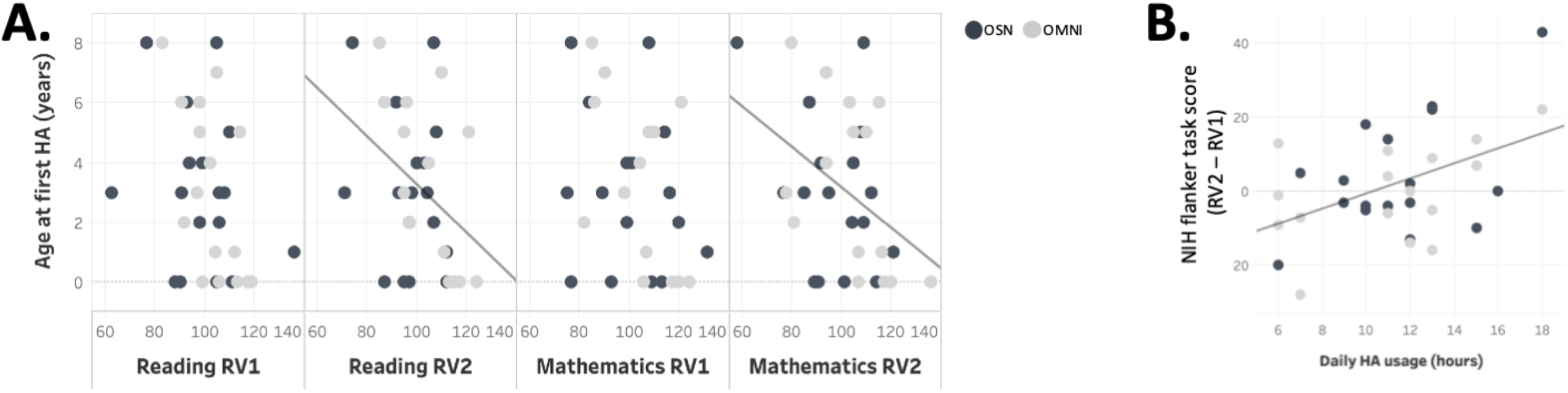
(A) Negative correlations between the children’s age at first hearing aid fit with WJ-IV Reading and Mathematics outcomes at RV2. (B) Positive correlation between daily hearing aid usage and change in performance on the NIH toolbox attention task (flanker task).

However, no other significant relationships between performance and age at first fit or duration of hearing aid use were found.

## DISCUSSION

Long term use of a hearing aid processing algorithm (OSN) designed to improve speech hearing in situations of competing speech neither enhanced nor reduced performance on a variety of age-standardized auditory, cognitive, and academic tasks, relative to a control, omnidirectional processor. Some changes in performance, both objective (perceptual, linguistic, cognitive, academic) and subjective (SSQ), were observed between RV1 and RV2 in both treatment groups. These were likely due to practice/reliability (Taylor et al, 2020) or caregiver expectation (SSQ). However, some exceptions to these otherwise unsurprising results were observed. One was the subjective improvement in spatial hearing of trial users only of the Omni processing strategy. Another was the emergence of a significant relationship between age at first hearing aid use and both Reading and Mathematics scores that became significant at RV2.

Enhanced target word repetition in SSN, compared with a two-talker masker (TTM), has previously been reported in children with mild/moderate hearing loss using OPN hearing aids, regardless of whether the target talker was directly in front of the child (Browning et al., 2019). Other studies using headphone stimulation have also shown more TTM in younger children, but not in older youth or adults (McCreery et al., 2020). In the study reported here, we confirmed these findings of greater TTM, and extended them by showing improved performance in all test conditions after 7-13 months of OPN hearing aid use. Browning and colleagues (2019) additionally found that performance using the OSN processing algorithm was superior to that of the same children using the Omni algorithm in SSN masking, but not in TTM. Here, different groups of children used OSN and Omni algorithms, and we did not find enhanced benefit of OSN during either SSN masking or TTM. It is possible that the difference between these studies was due to the reduced variance associated with retesting the same participants in two test conditions.

Previously, we found that word repetition in noise did not improve after 2-3 months of using OSN in a new fitting, but caregiver reports of ability improved over the study period (Pinkl et al., 2021). Here, we report improved word repetition in noise after 7-13 months of continuous use of OSN and Omni. In addition, we found a general improvement in caregiver-reported (SSQ) speech abilities, and improved spatial spatial ability in the Omni group. Together, these results provide evidence for longer-term acclimatization. Acclimatization may reflect enhanced training, motivation and/or expectation, along different timelines. For example, there may be a general and rapid, positive motivation and caregiver expectation after a new intervention (Gilliver et al., 2013), followed by a more gradual perceptual learning.

There is a growing literature on the effects of complex processing enhancements on speech perception using hearing aids (Cox et al., 2014; directional microphones - Magnusson et al., 2013; noise reduction - Brons et al., 2014; Magnusson et al., 2013; frequency compression - Hopkins et al., 2014; Picou et al., 2015). In a recent review, Lesica (2018) summed up these enhancements as showing only “modest additional benefit” (p. 175) beyond amplification. By showing no significant difference in a highly controlled study of the auditory and related cognitive and academic performance of children using omni or directional microphones with noise reduction processing strategies, the results reported here appear to support Lesica’s thesis. However, his optimistic suggestion was that alternative strategies based on recent developments in physiology and artificial intelligence should improve matters in the near future. In particular, an emphasis on the importance of cognitive and environmental factors to determine optimal, individualized processing strategies seems particularly suited to a pediatric population.

Moeller & Tomblin (2015) and Nakeva von Mentzer et al. (2020) have shown the importance of early and prompt fitting of hearing aids for children with hearing loss. The average age of our participants was 9 years 9 months and the length of hearing aid usage prior to the study was 4 years 1 month. We showed that, regardless of processing strategy, the age of hearing aid fit continued to have an effect on academic achievement during development with the relationship between age of first hearing aid use and reading/mathematics scores showing significance at RV2, and trending, but not surviving family correction at RV1.

The end of our study was conducted during the COVID-19 pandemic with eight of our participants (OSN = 5, omni = 3) completing RV2 during periods of school closures. It is of note that four of these children (OSN = 2, omni = 2) were not included in the final analysis as they stopped wearing their hearing aids consistently with their daily usage below 6 hours. Consistent daily use of HAs has been highlighted as a strong predictor of cognitive and academic scores (Walker et al., 2015b). We echo this with a positive significant relationship between daily HA use and change in performance on the selective attention (flanker) task. Adding a covariate of whether the participants completed testing during the pandemic did not affect our findings.

We have provided a broad spectrum of test approaches including auditory sensitivity, several forms of speech listening, learning in noise and in quiet, cognitive and academic performance, and caregiver- and self-report. On none of these tests was any significant performance difference found between the Omni and the OSN processing strategies. However, it remains a possibility that in more realistic test environments differences would be found between these strategies, for example, where distracting sounds were presented simultaneously from several different directions or where visual information is combined with auditory. While a within-subject design reduces individual differences, our study had multiple levels of quality checks built in. These included groups well matched in hearing level, audiogram shape and age, double-blinding throughout the study until the last participant completed their RV2, checklists to ensure all audiologists followed the same rigorous fitting protocols, and study pre-registration. We found no evidence that children learned how to use the additional features provided by OSN.

## Acknowledgments

HJS designed the protocol, registered the study, performed experiments, analyzed data and wrote the manuscript. EKC and JP performed experiments, analyzed data and wrote the manuscript. JP also registered the study. CNvM designed the protocol and commented on the manuscript. CCHMC Division of Audiology designed the protocol and performed experiments. LLH designed the protocol and commented on the manuscript. DRM designed the protocol and wrote the manuscript. All authors discussed the results and implications.

The authors would like to thank the large team involved in completing this study. Including Jasmin Martinez and Sarah Ferguson for behavioural data collection; the study audiologists Tommy Evans, Tim Nejman, Jeanie Hamilton, Anne-Marie Wollet and Erin Stewart; the MRI techs Matt Lanier, Lacey Haas, Kaley Bridgewater, Kelsey Murphy, Brynne Williams and Marty Jones; Audrey Perdew for database management; Andrew Wagner for programming support; Cecilia Nakeva von Mentzer for providing the sentence repetition task along with task interpretation; Boys Town National Research Hospital for supplying the word repetition in noise test stimuli; Li Lin for her assistance with statistical analyses; and Elaine Ng and Thomas Behrens from Oticon for help with funding, providing hearing aids, and other general support.

A preprint of this paper can be found at: medxriv

